# Comparability of venous and capillary blood measurements for a host-protein test differentiating bacterial from viral infections

**DOI:** 10.1101/2025.07.21.25331877

**Authors:** Mary Hainrichson, Nitzan Shamir, Naftalie Senderovich, Husein Darawsha, Ayelet Raz, Salim Halabi, Oded Shaham, Roy Navon, Tanya M. Gottlieb, Barak Hershkovitz

**Affiliations:** MeMed, 7 Nahum Het St, Tirat Carmel, Israel 3508506; Lin Medical Center, Rothschild Blvd 37, Haifa, Israel 3515210; Rambam Health Care Campus, HaAliya HaShniya St 8, Haifa, Israel 3109601; Carmel Medical Center, Mikhal St 7, Haifa, Israel 3436212

**Keywords:** Rapid host-protein test, capillary blood, TRAIL, IP-10 and CRP, viral, bacterial, MeMed BV, MeMed Key

## Abstract

**Background:** MeMed BV (MMBV) is a host-protein test for discriminating between bacterial and viral infections. It was analytically and clinically validated in serum and whole blood samples. Here we investigated the comparability of venous versus capillary blood measurements of the MeMed BV score and of the composite TRAIL, IP-10 and CRP biomarkers.

**Methods:** A prospective cohort study enrolled adult patients presenting with suspected acute infection between Oct-2024 till Jan-2025 at three medical centers. Eligibility criteria were according to the MeMed BV’s instructions for use. Paired venous and capillary blood specimens were collected from each patient. Each sample (50 µL) was measured on MeMed Key® (MeMed, US) with a next generation MeMed BV cartridge. Passing-Bablok regression was used to evaluate the relationship between venous and capillary measurements for each biomarker and the MeMed BV score. For the MeMed BV score, the first accuracy requirement was that the slope should fall in the range 0.9-1.1 and the intercept in the range of −5 to 5. The second accuracy requirement was that the 95% confidence intervals for the bias at all score bin cutoffs should fall within the interval [−12.5,12.5]. The clinical acceptance criterion for the score was that less than 5% of the paired sample measurements should fall into non-adjacent score bins.

**Results:** The study population comprised 58 patients with median age 58.5 (interquartile range: 36.25-75.75). Venous versus capillary blood measurements for TRAIL, IP-10 and CRP demonstrated high correlation with coefficients of 0.98, 0.98 and 0.99 respectively. For the MeMed BV score, the slope in the Passing-Bablok regression analysis was 1.00 (95% confidence interval, CI: 0.99 – 1.00) and the intercept 0.00 (0.00 – 0.08), satisfying the first accuracy requirement. The estimate of bias at each cutoff satisfied the second accuracy requirement: score 10, −0.57 (95%CI: −3.00 – 0.56); score 35, −0.47 (−2.10 – 0.38); score 65, −0.34 (−1.28 – 0.22); and score 90, −0.24 (−0.76 – 0.44). None of the paired samples generated MeMed BV scores falling into non-adjacent bins, satisfying the clinical criterion.

**Conclusion:** TRAIL, IP-10, CRP and MeMed BV score measurements are comparable in venous and capillary blood when performed using the next generation MeMed BV cartridge and MeMed Key. The present findings can serve as the basis for expanding MeMed BV’s use to include capillary blood specimens, potentially widening its clinical utility.

## Introduction

Accurate and timely identification of infection etiology is essential to guiding appropriate clinical care. Similarity of presenting signs and symptoms means it is often hard to discriminate between bacterial and viral infections, leading to diagnostic uncertainty and sub-optimal patient outcomes at Emergency Departments (EDs),^1^ Urgent Care Clinics (UCCs)^2^ and in primary care settings.^3^

Advanced host-response tools that leverage machine learning to combine multiple biomarkers into a bacterial likelihood score have emerged as promising adjunctive tests for determining infection etiology.^4^ A barrier to their wide adoption in decentralized settings and for the elderly and pediatric populations is the requirement for venous blood, which necessitates blood draw by a phlebotomist. To address this, capillary blood can be utilized, but the potential impact of employing a smaller blood volume and of composition differences in capillary versus venous blood need to be investigated. For example, capillary samples can contain interstitial fluid and exhibit hematocrit-related variability, potentially impacting biomarker quantification.^5^

MeMed BV is a host-protein test that integrates circulating levels of tumor necrosis factor-related apoptosis-inducing ligand (TRAIL), interferon gamma-induced protein-10 (IP-10), and C-reactive protein (CRP), to generate a bacterial likelihood score.^6^ MeMed BV was analytically validated in serum (100 ul) and whole blood (150 ul) samples.^7,8^ The test was clinically validated in children and adults in EDs and UCCs^9–17^ and its clinical utility is supported by real world^2,18^ and randomized control evidence.^1^ Here we investigate the comparability of venous versus capillary blood measurements of the MeMed BV score and of the composite TRAIL, IP-10 and CRP biomarkers.

## Materials and Methods

### Study Design and Population

A prospective cohort study was conducted to establish the comparability of the MeMed BV score and the levels of TRAIL, IP-10 and CRP measured in venous versus capillary whole blood specimens. Adult patients presenting with suspected acute infection were recruited between 30-Oct-2024 till 14-Jan-2025 at three medical centers. The study was approved by the ethics committees of each participating center and informed consent was required: Lin Clinic, Clalit Health Services, Haifa (IRB 0001-24-COM1), Rambam Health Care Campus (IRB 0285-17-RMB), Carmel Medical Center (IRB 0031-21-CMC).

Eligibility criteria were according to the MeMed BV’s instructions for use (MeMed, US). Briefly, the main inclusion criteria were symptoms of acute infectious disease and experienced fever within the last 7 days. Additionally, inclusion required successful sampling of paired venous blood and capillary blood specimens. The main exclusion criteria were active inflammatory disease, congenital or acquired immunodeficiency, chronic fungal or parasitic infection, hepatitis B virus, hepatitis C virus, infection with active tuberculosis, significant trauma or burns in the last 7 days, major surgery within the last 7 days, pregnancy and active malignancy. Patients were excluded if the blood clotted during capillary sampling.

### Sample Handling and Analysis

Paired venous and capillary blood specimens were collected from each patient. Venous blood was collected into 6 mL K_2_EDTA Vacutainer® tubes (BD, REF: 367864). Capillary blood was obtained via fingerstick into K_2_EDTA Microtainer® tubes (BD, REF: 365975). The samples were transferred at room temperature to the MeMed laboratory for testing within 90 minutes. Each sample (50 µL) was measured on MeMed Key® (MeMed, US) with a next generation MeMed BV cartridge. The paired venous and capillary samples were run for each patient. Each venous sample was run on three MeMed Key analyzers. Depending on the volume of the capillary blood available, the sample was run on three analyzers (n=24), or two (n=16) or one analyzer (n=18).

### MeMed BV test

The MeMed BV (MMBV) result is a score, ranging from 0 to 100, that computationally integrates TRAIL, IP-10, and CRP measurements using a previously derived algorithm^6^ which was employed in multiple previous studies.^9–11,13–15^ Higher scores are indicative of bacterial infection (or co-infection). In line with the manufacturer’s instructions for use, five MMBV score bins^6,9–11,13–15,17^ were employed for clinical interpretation:

- Bin 1: 0≤score≤10, high likelihood of viral infection (or other non-bacterial etiology)
- Bin 2: 10<score<35, moderate likelihood of viral infection (or other non-bacterial etiology)
- Bin 3: 35≤score≤65, equivocal
- Bin 4: 65<score<90, moderate likelihood of bacterial infection (including coinfection)
- Bin 5: 90≤score≤100, high likelihood of bacterial infection (including coinfection).

### Statistical Approach

Comparability between venous and capillary blood samples was established based on CLSI guideline EP09: “Measurement procedure comparison and bias estimation” and EP35: “Assessment of Equivalence or suitability of specimen types”. The mean biomarker or MeMed BV score measurement for each sample was averaged from the values generated by the 1-3 MeMed Key analyzers.

For the individual biomarkers, comparability between venous and capillary blood samples was demonstrated by conducting Passing-Bablok regression. There was no accuracy criterion for the individual biomarkers.

For the MeMed BV score, the accuracy requirement was that the slope should fall in the range 0.9-1.1 and the intercept in the range of −5 to 5 of the Passing-Bablok regression analysis.

Additionally, the bias based on Deming regression analysis incurred by using capillary blood as compared to venous blood was calculated at the bin cutoffs (10, 35, 65, 90), as described previously.^7^ The ratio of error variances was set at λ=1. The second criterion for accuracy was that the 95% confidence intervals for the bias at all score bin cutoff should fall within the interval [−12.5,12.5].

For the MeMed BV score there was also a clinical acceptance criterion: the mean score measured using capillary blood should deviate from the corresponding mean score measured using venous blood by an amount that would place the pair of scores in two non-adjacent bins in less than 5% of the samples.

## Results

The eligible study population comprised 58 patients with median age 58.5 years (interquartile range, IQR: 36.25-75.75).

### TRAIL, IP-10 and CRP in capillary versus venous blood

For each biomarker, measurements generated using venous blood were compared to those generated using capillary blood (Figure 1 and Table 1). To be conservative, the venous blood samples with biomarker measurements below the lower limit of quantitation (LoQ) or above the higher LoQ were removed from the regression analysis for the relevant biomarker.

**Table 1.**
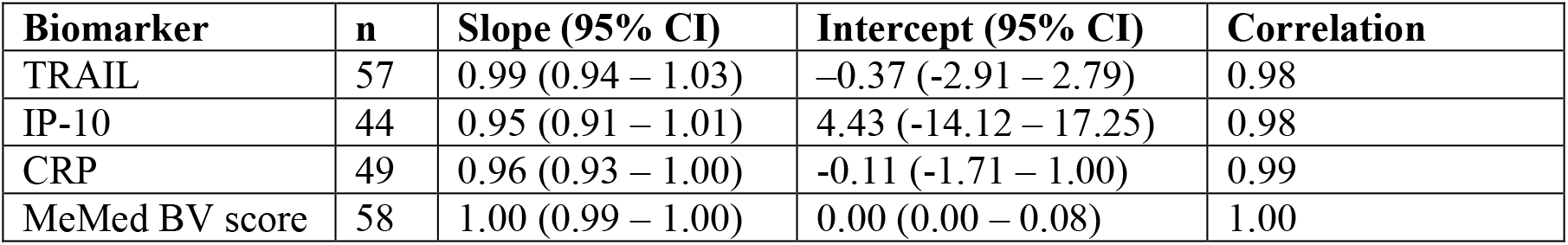
Slope, intercept and correlation coefficient values for Passing-Bablok regression analysis.

**Figure 1.**
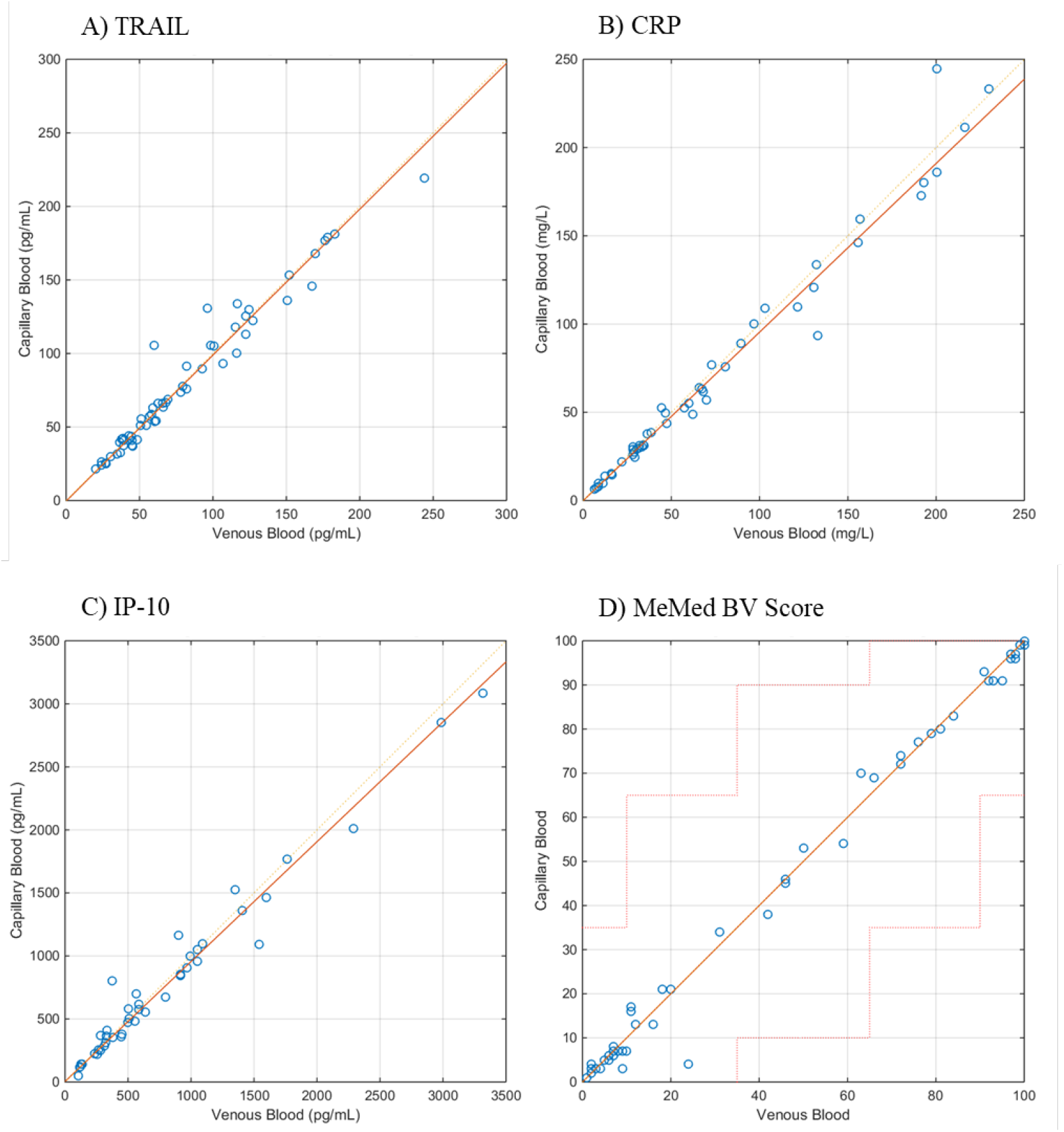
Correlation between measurements in venous (x-axis) versus capillary (y-axis) blood for TRAIL (panel A), CRP (panel B), IP-10 (panel C) and MeMed BV score (panel D). For the biomarkers, the red line represents the regression line, and the red dash line represents the identity line. For the BV score, the red line represents the regression line, and the red dash line represents the limit to pass to a non-adjacent bin.

### MeMed BV score in capillary versus venous blood

For the MeMed BV result, scores generated using venous blood were compared to those generated using capillary blood (Figure 1). The accuracy criteria for slope and intercept were satisfied (Table1). The estimate of bias at each cutoff was: score 10, −0.57 (95% confidence interval, CI −3.00 – 0.56); score 35, −0.47 (−2.10 – 0.38); score 65, −0.34 (−1.28 – 0.22); and score 90, −0.24 (−0.76 – 0.44). Accordingly, the second accuracy criterion was satisfied.

There were zero samples with pairs of scores falling in non-adjacent bins, satisfying the clinical criterion.

## Discussion

This study established comparability between venous and capillary blood (50 µL) measurements for TRAIL, IP-10, CRP and MeMed BV score using the MeMed Key and next generation MeMed BV test. These results align with a previous study reporting the equivalency of CRP measurements in capillary versus venous blood.^19^

Study limitations include the controlled laboratory conditions, which may not reflect real world variability in sample collection and handling. Study strengths include the use of routinely available, off-the-shelf blood collection tubes, enabling replicability and translation into practice.

Multiple reports support that capillary blood measurements are less invasive and more practicable in outpatient and pediatric settings, due to logistical and patient-centered benefits.^20,21^ The present findings can serve as the basis for expanding MeMed BV’s use to include capillary blood specimens, potentially widening its clinical utility. Further studies are warranted to examine comparability in real world settings where samples may be collected by users with variable proficiency.

## Data Availability

All data produced in the present study are available upon reasonable request to the authors.

## Abbreviations

MMBV: MeMed BV
TRAIL TNF: TNF related apoptosis-inducing ligand
IP-10: interferon-gamma inducible protein of 10 kDa
CRP: C-reactive protein
IQR: interquartile range
UCC: urgent care center
ED: emergency department
LoQ: limit of quantitation
CI: confidence interval.

## Acknowledgements

We thank our colleagues for their inputs: Nataly Agra, Einav Simon, Lior Kellerman and Eran Eden.

